# Initiation of buprenorphine as part of pain management approach to trauma patients in the intensive care unit with a history of opioid use disorder: A QI Study

**DOI:** 10.64898/2026.03.06.26347723

**Authors:** Abdurrehman Khan, Bedda Rosario-Rivera, Sharvari P. Shivanekar, Jason L. Sperry, Trent D. Emerick, Ata Murat Kaynar

**Affiliations:** Department of Anesthesiology and Perioperative Medicine, University of Pittsburgh in Pittsburgh, Pittsburgh, PA 15206; Department of Epidemiology, University of Pittsburgh in Pittsburgh, Pittsburgh, PA 15206; Department of Psychiatry, University of Pittsburgh in Pittsburgh, Pittsburgh, PA 15206; Department of Critical Care Medicine, University of Pittsburgh in Pittsburgh, Pittsburgh, PA 15206; Department of Surgery, University of Pittsburgh; Pittsburgh, PA 15213, USA; Division of Trauma and Acute Care Surgery, Pittsburgh Trauma and Transfusion Medicine Research Center, Pittsburgh, PA 15213, USA; The Clinical Research, Investigation, and Systems Modeling of Acute Illness (CRISMA) Center, University of Pittsburgh in Pittsburgh, Pittsburgh, PA 15206; The Center for Innovation in Pain Care (CIPC), University of Pittsburgh in Pittsburgh, Pittsburgh, PA 15206

**Keywords:** Buprenorphine, ICU, acute pain management

## Abstract

**Background:** Buprenorphine use in the intensive care unit (ICU) remains not well studied despite growing perioperative guidance supporting its continuation and initiation for patients with opioid use disorder (OUD). Trauma ICU admissions represent a critical opportunity to address untreated OUD, as well as continuation of an already established treatment plan for OUD, yet barriers limit its adoption in this setting.

**Methods:** This single-center quality improvement study evaluated for inpatient buprenorphine prescribing patterns following provider education at a tertiary academic trauma center. Adult trauma ICU patients with OUD admitted between 2016–2024 were identified through the institutional trauma registry. Patients with pre-admission buprenorphine were excluded, yielding a cohort of 95 patients: 24 buprenorphine-exposed (initiated in the hospital) and 71 controls. Primary outcomes included pain scores and opioid requirements (morphine milligram equivalents, MME) during the first 48 hours. Secondary outcomes included hospital length of stay (LOS), discharge disposition, and 90-day readmission.

**Results:** Baseline characteristics were similar between groups. No statistically significant differences were observed in first recorded pain scores (median 8 vs. 10; p=0.35), mean 48-hour pain scores (7.40 vs. 7.76; p=0.44), or total opioid consumption (232 vs. 119 mg MME; p=0.45). Median hospital LOS (16 vs. 19 days; p=0.48) and 90-day readmission rates (42.3% vs. 33.3%; p=0.40) were also comparable.

**Conclusion:** Inpatient buprenorphine initiation in trauma ICU patients with OUD was not associated with worse pain control, increased opioid requirements, or adverse clinical outcomes. These findings support the integration of buprenorphine into critical care pathways as a safe strategy to address OUD during hospitalization and improve long-term recovery continuity.

## Introduction

Guidelines for buprenorphine utilization in the perioperative period have become more established over the last five years; however, buprenorphine’s role in the intensive care unit (ICU) remains understudied.^1^ The initiation or maintenance of buprenorphine offers a pragmatic bridge between medication assisted treatment (MAT) and analgesia for trauma patients with opioid use disorder (OUD), encountering acute injury, surgery, and risk for withdrawal, relapse, and overdose if management is fragmented.^2^ Contemporary perioperative guidance advises against routine discontinuation and supports in-hospital buprenorphine initiation when OUD is newly identified-an approach that stabilize cravings, enable multimodal analgesia, and possibly reducing perioperative complications.^3^ As a high-affinity partial µ-agonist/κ-antagonist, buprenorphine prevents withdrawal in opioid tolerant patients while providing analgesia; strategies that lower reliance on full µ-agonists may mitigate misuse risks.^4^

Practical perioperative guidelines offer frameworks for buprenorphine management alongside regional and non-opioid analgesics.^5^ However, implementation barriers persist, such as clinician preparedness and lack of experience treating OUD.^5^ This quality improvement (QI) study was conducted to examine trends in ICU buprenorphine prescribing following provider education. The analysis focused on buprenorphine formulations FDA-approved for OUD treatment rather than those indicated for pain.

## Methods

### Study Design and Setting

We conducted this QI study at a tertiary academic trauma center following provider education to evaluate buprenorphine prescription and associated clinical outcomes among hospitalized patients with OUD admitted to the trauma ICU. This study was approved by UPMC Presbyterian Quality Improvement Committee #4966.

### Study Population

We identified adult trauma ICU patients (≥18 years) with OUD admitted between 2016 and 2024 through the institutional trauma registry. We included 109 unique patients. Buprenorphine exposure was determined using linked outpatient prescription records with inpatient medication administrative data. Buprenorphine exposure was defined as receipt of ≥1 inpatient buprenorphine dose during hospitalization (Table 1). Patients were grouped into a buprenorphine-exposed group and a control group with no inpatient exposure. Patients who received buprenorphine prior to admission (n=14) were excluded, leaving a cohort of 95 patients (71 control, 24 buprenorphine-exposed). Buprenorphine formulations included sublingual film and tablet forms that are approved for OUD.

**Table 1.** Descriptive statistics for buprenorphine forms.

| **Buprenorphine medication form** | **Frequency (%)** |
| --- | --- |
| Suboxone 2 mg-0.5 mg sublingual tablet, disintegrating | 16 (1.5) |
| Suboxone 2mg-0.5mg sublingual film | 190 (18.1) |
| Suboxone 8 mg-2 mg sublingual tablet, disintegrating | 120 (11.4) |
| Suboxone 8mg-2mg sublingual film | 605 (57.6) |
| Subutex | 1 (0.1) |
| Buprenorphine (generic sublingual) | 111 (10.6) |
| buprenorphine-naloxone (Suboxone) 2mg-0.5mg sublingual film | 1 (0.1) |
| buprenorphine-naloxone (Suboxone) 8mg-2mg sublingual film | 7 (0.7) |

### Outcomes

Primary outcome: Pain scores and opioid requirements during the first 48 hours of ICU care. Pain scores (0–10 Numeric Rating Scale) were extracted from nursing documentation; implausible entries and repeated timestamps were removed. Pain outcomes included the first recorded score within 48 hours and the mean score at 24–48 hours. Total opioid exposure was converted to morphine milligram equivalents (MME). Secondary outcomes included hospital length of stay (LOS), discharge disposition, and 90-day readmission.

### Statistical Analysis

Demographics and outcomes were summarized and reported as mean (standard deviations), median (interquartile range, IQR) or frequency (percentages). Distribution assumptions for continuous data were evaluated using histograms and Q-Q plots. Continuous variables were compared between groups using the Wilcoxon rank-sum test, and categorical variables using Chi-square or Fisher’s exact test. A negative binomial model was used to compare hospital LOS between the two groups due to right-skewed data and overdispersion. Statistical significance was defined as p<0.05. All statistical analyses were conducted using SAS 9.4 (SAS Institute, Cary, NC).

## Results

A total of 95 trauma ICU patients with OUD, who underwent a surgical procedure, were included, of whom 24 (25.3%) received inpatient buprenorphine and 71 (74.7%) did not. Baseline demographic and clinical characteristics were similar between groups (Table 2).

**Table 2.** Descriptive statistics for baseline characteristics and outcomes.

|  |  | N | All | N | Control | N | Buprenorphine | p |
| --- | --- | --- | --- | --- | --- | --- | --- | --- |
| **Demographic/clinical characteristic** | **Label** |  |  |  |  |  |  |  |
| **Age(y)** |  | 95 | 47.5<br>±<br>13.5 | 71 | 48.4 ±<br>14.2 | 24 | 44.9 ± 10.9 | 0.27 |
| **Sex** |  | 95 |  | 71 |  | 24 |  | 0.11 |
|  | **Female** |  | 45<br>(47.4) |  | 37<br>(52.1) |  | 8 (33.3) |  |
|  | **Male** |  | 50<br>(52.6) |  | 34<br>(47.9) |  | 16 (66.7) |  |
| **Race** |  | 95 |  | 71 |  | 24 |  | 0.36* |
|  | **White** |  | 74<br>(77.9) |  | 53<br>(74.6) |  | 21 (87.5) |  |
|  | **Black** |  | 16<br>(16.8) |  | 13<br>(18.3) |  | 3 (12.5) |  |
|  | **Declined** |  | 5<br>(5.3) |  | 5 (7.0) |  | 0 (0.0) |  |
| **Ethnicity** |  | 95 |  | 71 |  | 24 |  | 0.05 |
|  | **Non-Hispanic** |  | 73<br>(76.8) |  | 58<br>(81.7) |  | 15 (62.5) |  |
|  | **Unknown** |  | 22<br>(23.2) |  | 13<br>(18.3) |  | 9 (37.5) |  |
| **ASA Class** |  | 95 |  | 71 |  | 24 |  | 0.14* |
|  | **2** |  | 2<br>(2.1) |  | 0 (0.0) |  | 2 (8.3) |  |
|  | **3** |  | 19<br>(20.0) |  | 12<br>(16.9) |  | 7 (29.2) |  |
|  | **3E** |  | 20<br>(21.1) |  | 17<br>(23.9) |  | 3 (12.5) |  |
|  | **4** |  | 13<br>(13.7) |  | 10<br>(14.1) |  | 3 (12.5) |  |
|  | **4E** |  | 38<br>(40.0) |  | 30<br>(42.3) |  | 8 (33.3) |  |
|  | **5E** |  | 3<br>(3.2) |  | 2 (2.8) |  | 1 (4.2) |  |
| **Disposition** |  | 95 |  | 71 |  | 24 |  | 0.83* |
|  | **Home** |  | 42<br>(44.2) |  | 30<br>(42.3) |  | 12 (50.0) |  |
|  | **Hospice** |  | 4<br>(4.2) |  | 3 (4.2) |  | 1 (4.2) |  |
|  | **LTACH/Rehab** |  | 18<br>(18.9) |  | 13<br>(18.3) |  | 5 (20.8) |  |
|  | **Other** |  | 31<br>(32.6) |  | 25<br>(35.2) |  | 6 (25.0) |  |
| **Patient Priority** | **Patient Priority** | 95 |  | 71 |  | 24 |  | 0.57* |
|  | **Trauma Center** |  | 37<br>(38.9) |  | 25<br>(35.2) |  | 12 (50.0) |  |
|  | **Emergency** |  | 42 |  | 34 |  | 8 (33.3) |  |
|  | **Dept** |  | (44.2) |  | (47.9) |  |  |  |
|  | **Urgent** |  | 12<br>(12.6) |  | 9 (12.7) |  | 3 (12.5) |  |
|  | **Other** |  | 4<br>(4.2) |  | 3 (4.2) |  | 1 (4.2) |  |
| **Outcomes** |  |  |  |  |  |  |  |  |
| **First Pain Score Prior Procedure** |  | 62 | 9 [7-10] | 46 | 8 [7-10] | 16 | 9.5 [7.75-10] |  |
| **First Pain Score After Procedure** |  | 64 | 8.5 [6-10] | 45 | 8 [6-10] | 19 | 10 [6-10] |  |
| **Average 48hr Pain Score** |  | 64 | 7.63 [6.61-8.78] | 45 | 7.4 [6.55-8.73] | 19 | 7.76 [6.68-8.94] |  |
| **Average 24-48hr Pain Score** |  | 59 | 7.68 [7-8.89] | 42 | 7.58 [7-8.67] | 17 | 8 [7.44-9] |  |
| **Total MME within 48hr** |  | 95 | 185 [43.4-648] | 71 | 232 [43.4-793] | 24 | 119 [42.9-509] |  |
| **Total MME within 48hr (outliers removed)** |  | 80 |  | 58 | 89.2 [30.9-393] | 22 | 103 [31.7-402] |  |
| **Total MME within 24-48hr** |  | 95 | 58.4 [15.9-389] | 71 | 88.2 [17.5-443] | 24 | 49.1 [14.2-223] |  |
| **Total MME within 24-48hr (outliers removed)** |  | 78 |  | 56 | 35.0 [12.8-194.2] | 22 | 49.1 [15.0-1669.0] |  |
| **LOS Days** |  | 95 | 17 [9-28] | 71 | 16 [9-27] | 24 | 19 [10.5-36.5] |  |
| **Readmission within 90 days** |  | 95 | 38 (40.0) | 71 | 30 (42.3) | 24 | 8 (33.3) |  |

Mean ± standard deviation, median [interquartile range (IQR), first quartile – third quartile] or frequency (percent); ASA Class, American Society of Anesthesiologists classification; LTACH, long term acute care hospital; MME, morphine milligram equivalents; LOS, length of stay \*Fisher's exact test

Pain score data was only available for 64 patients postoperatively and 59 patients for the 24–48-hour analysis window due to incomplete documentation (Table 2). Outliers, defined as observations >1.5×IQR above the 75^th^ percentile, were removed for MME calculations, as well.

## Discussion

This QI study evaluated inpatient buprenorphine initiation among trauma ICU patients with OUD and found no statistically significant differences in pain scores, opioid requirements, hospital length of stay, or 90-day readmission compared to those without buprenorphine. These findings suggest that initiating buprenorphine during critical illness is not associated with worse in-hospital and out-of-hospital outcomes and may be incorporated into ICU care pathways for patients with OUD. This study enforces that early initiation of buprenorphine for patients with a history of trauma and OUD, even in the critical care unit, should be considered.

Although buprenorphine utilization in the perioperative period has become increasingly supported by expert guidance, its role in the ICU remains less clearly defined.^3,6^ In a recent study among ICU patients, sublingual buprenorphine was shown to be effective for analgesia.^1^ Our findings align with these observations by demonstrating that buprenorphine initiation was not associated with increased pain burden or higher opioid consumption in the ICU and additionally showing a trend of lower 90-day readmission trends.

Our study goal was to educate providers on the buprenorphine initiation for critically ill patients with OUD during the index inpatient admissions. Others have shown that in-patient buprenorphine initiation increases engagement in addiction treatment and reduces illicit opioid use and overdose risk following discharge.^7,8^ Similarly, patients initiated on buprenorphine during hospitalization had significantly higher treatment retention after discharge compared to those who received detoxification alone.^8^ Hospital encounters, particularly in critical illness, represent an opportunity to address untreated OUD and bridge patients to long-term treatment.^9^ However, clinician uncertainty regarding induction strategies, concerns about respiratory depression in opioid-tolerant patients, and the misconceptions that buprenorphine complicates analgesia are common barriers to its use in the ICU.^6,10^

This study has limitations inherent to retrospective, single-center analyses; missing pain assessments reduced sample size. Confounding may be present due to unmeasured differences in illness severity, injury burden, and baseline MMEs– which would be difficult to compare due to the use of pre-admission non-prescribed opioids in many cases– as no baseline covariate adjustment was performed. Additionally, this study was not powered to detect small differences in outcomes between groups. The timing of the first buprenorphine dose was also not controlled for.

Our findings, paired with other literature, suggest that initiating buprenorphine in the trauma ICU is safe and may support continuity of OUD care without compromising short-term clinical outcomes. Implementation of structured buprenorphine pathways in critical care settings may reduce missed opportunities to treat OUD during hospitalization and improve long-term recovery trajectories.

## Data Availability

All data produced in the present work are contained in the manuscript

